# Enhancing retention on antiretroviral therapy at 6 months using interactive two-way texting: findings from a randomized controlled trial in Lilongwe, Malawi

**DOI:** 10.1101/2024.11.02.24316629

**Authors:** Christine Kiruthu-Kamamia, Robin E. Klabbers, Hannock Tweya, Jacqueline Huwa, Agness Thawani, Pachawo Bisani, Joseph Chintedza, Geldert Chiwaya, Aubrey G. Kudzala, Dumisani Ndhlovu, Johnnie Seyani, Wim Groot, Milena Pavlova, Caryl Feldacker

## Abstract

Antiretroviral therapy (ART) retention is critical for achieving viral load suppression (VLS) among people living with HIV (PLHIV). Retention remains challenging in high-prevalence settings like Malawi. Short messaging service (SMS) interventions, particularly hybrid two-way texting (2wT), show promise in improving ART retention. We conducted a randomized control trial (RCT) at Lighthouse Trust in Lilongwe, Malawi, to evaluate the effectiveness of a hybrid 2wT system to improve early retention, appointment attendance, and VLS among new ART initiates within six months of ART initiation. After receiving routine ART initiation counseling, 452 new ART clients with mobile phones were randomized to 2wT or standard of care (SoC). The 2wT group received weekly motivational messages, appointment reminders, and had access to an open-ended SMS communication channel with healthcare workers. The SoC group received peer support at clinic visits and visit reminder phone calls. All participants were traced if they missed a clinic appointment by 14 days. Study outcomes included: retention in care (alive on ART), appointment adherence (attending within 2 days), and VLS (< 200 copies) at six months. Data from electronic medical records were analyzed using Chi-square tests and multivariable logistic regression. At six months post ART initiation, the 2wT group demonstrated significantly higher appointment adherence (59.6% vs. 46.8%, p = 0.008) and VLS (97.5% vs. 93.2%, p=0.007) compared to SoC. Among both 2wT and SoC, 91% remained in care (p=0.68). Although retention among 2wT and SoC at 6 months did not differ, 2wT clients were more likely to attend clinic visits on time and reach VLS in the first six months. The low-tech 2wT approach offers a scalable, appropriate intervention to enhance visit compliance and VLS among PLHIV with mobile phones. Implementing 2wT study over a longer time frame and among more clients would likely provide evidence for scaling 2wT more broadly.

## Background

Despite significant global progress in initiating people living with HIV (PLHIV) onto antiretroviral therapy (ART), retention remains a challenge, especially during the first 12 months of ART (1–3). Retention is pivotal in achieving viral load suppression (VLS) (4,5). Malawi, with an HIV prevalence rate of 6.7% among adults (6), is progressing toward the UNAIDS 95-95-95 targets: 88% of PLHIV are aware of their status, 98% of those PLHIV who are aware of their status are on ART, and 97% of PLHIV on ART achieved VLS in 2021(7). Still, challenges persist in retaining individuals in ART care over time (8,9). The use of short messaging services (SMS) shows promise to enhance ART adherence, provide appointment reminders, and facilitate communication with healthcare workers (10–13). Evidence suggests that interactive two-way messaging, which allows for messages with response options between clients and healthcare workers, is more effective than one-way informational messaging in improving ART-related care outcomes (10,14,15).

In 2021, the Lighthouse Trust Martin Preuss Center ART clinic in Malawi in collaboration with the International Training and Education Center for Health (I-TECH) at the University of Washington and technology partner Medic conducted a quasi-experimental study to design, develop, test, and assess an interactive two-way texting (2wT) system to improve early retention among new ART initiates (16–18). Lighthouse Trust implements an early retention Differentiated Service Delivery (DSD) model for new ART clients during their first 12 months on ART, providing targeted support during this critical period. In this DSD model, focused on newly initiated ART clients, 2wT demonstrated high usability and acceptability for both clients and healthcare workers(17,18), improved ART retention at 12 months by 10% (19), and appeared cost-effective when considered at scale(20).

To strengthen the quasi-experimental findings, we conducted a small, pragmatic, randomized controlled trial (RCT) among new ART clients with phones at Lighthouse Trust ART clinics in Lilongwe, Malawi, to evaluate the 2wT impact on appointment attendance, retention on ART, and VLS at 6-months. We hypothesized that 2wT clients would show higher retention, clinic attendance, and VLS at 6 months post-ART initiation compared to standard of care (SoC) peers.

## Methods

### Setting

Lighthouse Trust (LT), a WHO-recognized Center of Excellence for HIV care, has partnered with the Malawi Ministry of Health (MoH) since 2001 to operate public ART clinics (21–23). LT operates two clinics in Lilongwe: Lighthouse Clinic at Kamuzu Central Hospital and Martin Preuss Center at Bwaila Hospital. These clinics provide free HIV care to over 38,000 clients and start over 500 clients on ART quarterly. Newly diagnosed clients start ART the same day and are registered in the Electronic Medical Records System (EMRS). Clients receive monthly visits for six months, with fewer visits for stable clients. Viral load suppression (VLS) testing is done six months post-ART initiation and then annually thereafter. The EMRS tracks visits, medications, side effects, adherence, VLS results, and ART outcomes.

### Study design, recruitment, and enrollment

Participants in this two-arm, prospective RCT were recruited between December 16 2022 and June 28 2023. Eligibility criteria included recent ART initiation (within 2 months), age 18 or older, phone ownership, basic literacy, and verification of enrolment message. Consenting participants were randomized 1:1 to either the 2wT (intervention) or SoC (control) using a block design in groups of 10. To detect a 59% reduction in dropout hazard with 80% power, we required 394 participants (197 per arm). Accounting for potential 10% attrition, we aimed to enrol 500 participants: 467 participants were enrolled and followed for 6 months post-ART initiation. 2wT participants chose Chichewa or English messages; all SMS communication was free of charge. The study was registered with the Clinical Trial Registry on 9/6/2022 (NCT05531448). The Consolidated Standards of Reporting Trials (CONSORT)Outcomes 2022 checklist(24) was used in conducting the study (S1 Table 1).

**Table 1:** Participant Characteristics of clients enrolled in SoC and 2wT.

|  | SoC<br>N=228 |  | 2wT<br>N=214 |  | P-value |
| --- | --- | --- | --- | --- | --- |
|  | N | % | N | % |  |
| Study site |  |  |  |  |  |
| LH | 63 | 27.6% | 58 | 27.1% | 0.901 |
| MPC | 165 | 72.4% | 156 | 72.9% |  |
| Sex |  |  |  |  |  |
| Male | 118 | 51.8% | 98 | 45.8% | 0.21 |
| Female | 110 | 48.2% | 116 | 54.2% |  |
| Age ( Median [IQR]) | 35 | [28-42.5] | 36 | [29-42] |  |
| Age group |  |  |  |  |  |
| 18-19 | 6 | 2.6% | 3 | 1.4% | 0.59 |
| 20-24 | 22 | 9.6% | 17 | 7.9% |  |
| 25-29 | 39 | 17.1% | 42 | 19.6% |  |
| 30-34 | 42 | 18.4% | 34 | 15.9% |  |
| 35-39 | 46 | 20.2% | 37 | 17.3% |  |
| 40-44 | 31 | 13.6% | 42 | 19.6% |  |
| 45-49 | 21 | 9.2% | 23 | 10.7% |  |
| 50+ | 21 | 9.2% | 16 | 7.5% |  |
| WHO HIV stage |  |  |  |  |  |
| 1 or 2 | 167 | 74.2% | 140 | 66.4% | 0.085 |
| 3 | 43 | 19.1% | 45 | 21.3% |  |
| 4 | 15 | 6.7% | 26 | 12.3% |  |
| Average visits<br>scheduled (median<br>[IQR]) | 3 | [2.000-4.000] | 3 | [2.000-4.000] | 0.509 |
| Overall visit compliance |  |  |  |  |  |
| Yes | 103 | 46.8% | 127 | 59.6% | 0.008 |
| No | 117 | 53.20% | 86 | 40.40% |  |

LH=Lighthouse Trust; MPC=Martin Preuss Center; IQR=interquartile range; WHO=World Health Organization

### The study interventions

#### Standard of care (SoC)

SoC clients received early retention support through expert client (EC) buddies and late retention support through the Back to Care (B2C) tracing program. For the first year on ART, SoC clients are supported by an EC buddy who is a PLHIV. EC buddies offer in-clinic ART adherence counseling, call clients for appointment reminders, and follow-ups with phone calls within 14 days of a client missing an appointment (19,20). Fourteen days after a missed visit, all LT clients at both facilities are referred to the B2C tracing program, where the B2C team calls or visits clients to return them to care (25).

#### Two-way texting (2wT) intervention

The 2wT system, detailed previously (17–20), was designed as an alternative to EC buddy support which is labor and time-intensive. 2wT clients received automated, weekly motivational messages, individualized SMS appointment reminders three days and one day before a scheduled appointment, and SMS reminders (if needed) 2, 5, and 11 days after a missed appointment. Additionally, 2wT accommodates open-ended SMS communication between clients and healthcare workers (HCWs) to reschedule visits, report transfers, or discuss non-clinical issues related to their ART care. Currently, 2wT clients who do not stop messages will continue 2wT support indefinitely. 2wT clients are also referred to B2C 14 days after a missed appointment, receiving the same return-to-care support as SoC clients. 2wT clients do not receive EC buddy support. Those who opted out of 2wT after receiving 2wT services were classified as withdrawn.

## Data Collection

Data on MoH ART outcomes, VLS, and client visits at six months post-ART initiation for both the intervention and control groups were extracted from the EMRS. ART outcomes include: alive in care (individuals retained in ART care as of the date of record review), stopped ART treatment (notified the clinic of their decision to stop treatment), transferred out (documented relocation), died (death due to any cause), and lost to follow-up (LTFU) (did not return to the clinic within 60 days of a scheduled visit)(26).

## Study outcomes

Study outcomes measured at 6 months post-ART initiation included: 1) Proportion of clients retained in care (alive in care); (2) adherence to clinic visit appointments (attended within 2 days of the scheduled appointment); and 3) VLS (suppressed = HIV RNA < 200 copies).

## Statistical Analysis

2wT participants were classified as exposed to 2wT and included in the analysis if they received at least one reminder SMS during the 6-month follow-up. Descriptive statistics compared participant characteristics as well as overall clinic visit compliance. The distribution of MoH ART outcomes 6 months post-ART initiation was compared using Chi-square tests to compare the distribution of on-time clinic visit compliance and VLS. A multivariable logistic regression analysis was used to estimate the odds of achieving VLS, adjusting for age, sex, intervention arm, ART visit compliance, and WHO stage at initiation. The intervention effectiveness on retention was assessed using time-to-event analysis, with transfer outs and death considered censoring events. LTFU and stopped ART treatment were considered failure events. Participants were censored 6 months after ART initiation, accounting for accrued time on ART for those joining after ART initiation. Kaplan-Meier plots displayed retention rates, comparing 2wT and control groups using log-rank tests. Participants who withdrew from 2wT were excluded.

## Ethical Consideration

The study protocol received approval from both the Malawi National Health Sciences Research Committee (#22/10/3036) and the University of Washington, Seattle, USA, ethics review board (STUDY00010106). All participants provided written informed consent in either Chichewa or English before enrolling in the study.

## Results

Included and excluded clients and the reasons for their exclusion are represented in the consort flow chart (S2 Fig 1). Clients were evenly distributed between clinics (Table 1). About half of all participants were female; most were WHO clinical stages 1 or 2 at ART initiation; and the median of scheduled visits during the study was 3 (IQR 2.000-4.000; p = 0.509). Overall visit compliance was significantly higher in the 2wT group, with 59.6% of participants adhering to scheduled visits compared to 46.8% in the SoC group (p = 0.008).

Six months after ART initiation, 91.1% of 2wT participants were alive and in care, compared to 91.2% in SoC (Table 2). Compared to SoC, 2wT had a lower proportion of LTFU (2.8% vs. 3.9%) and a higher proportion of clients who transferred out (4.2% vs. 3.5%) and died (1.9% vs. 0.9%) at six months, but the differences were not statistically significant (p = 0.68). In both groups, the majority achieved VLS at six months, with higher VLS among 2wT (97.5%) compared to SoC (93.2%) (p=0.007).

**Table 2:**
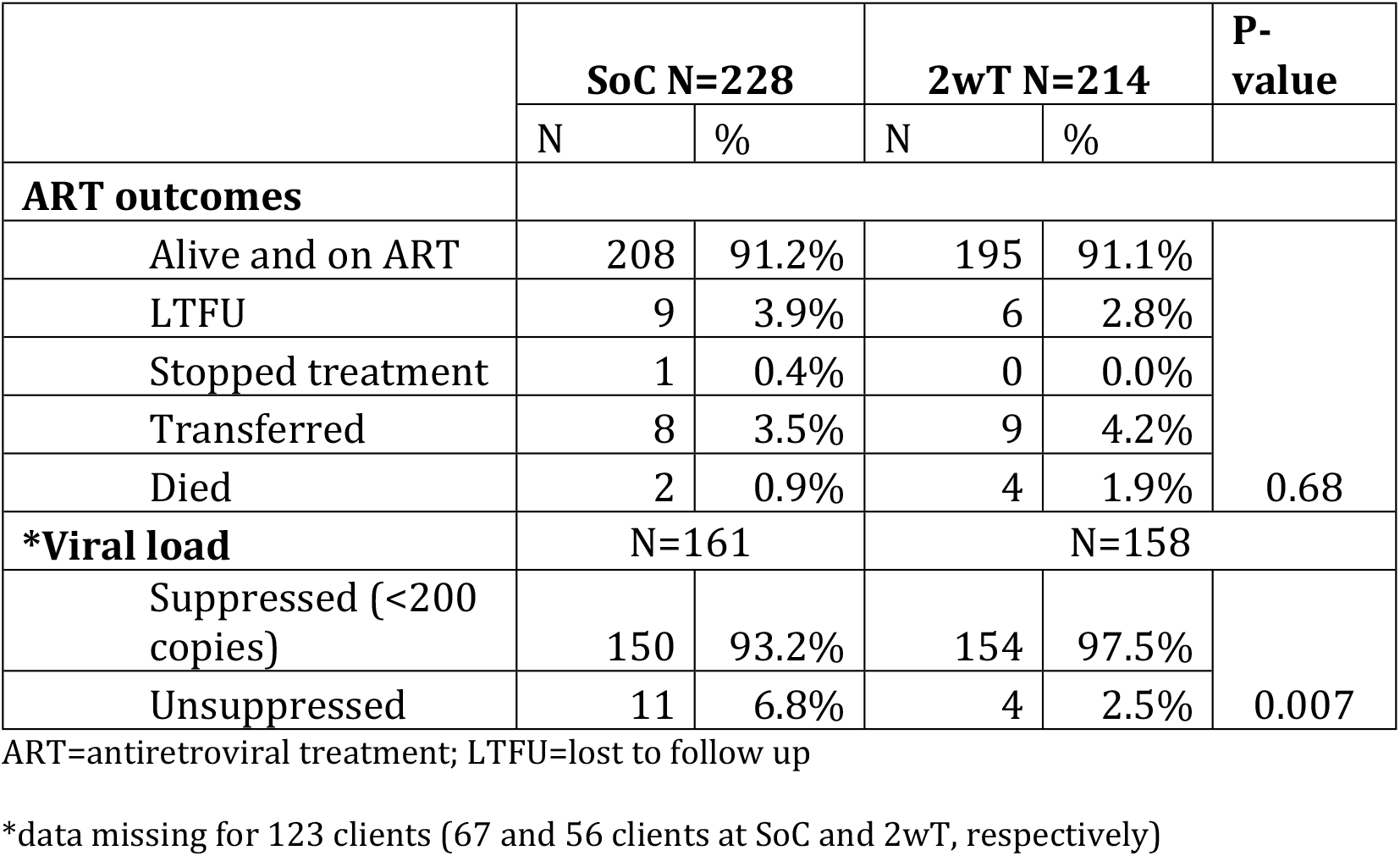
Association of Intervention Arm with ART Outcomes and Viral Load Suppression.

In the adjusted logistic regression analysis controlling for age, sex, intervention arm, ART visit compliance, and WHO stage (Table 3), the odds of achieving VLS was 2.74 times higher in 2wT clients than those in the SoC (OR = 2.74, 95% CI: 0.83 - 9.03, p = 0.098) albeit marginally (Table 3). Similarly, those who maintained overall on-time visit compliance (OR = 2.60, 95% CI: 0.84 - 8.01, p = 0.097) and women (OR = 0.34, 95% CI: 0.10 - 1.17, p = 0.086) were more likely to achieve VLS. Age (35+) and WHO stage at enrollment did not affect VLS.

**Table 3:** Logistic Regression Analysis of Achieving Viral Load Suppression.

| **Variable** | **Odds Ratio\*** | **p-value** | **95% Confidence Interval** |
| --- | --- | --- | --- |
| Intervention Arm | 2.7402 | 0.098 | 0.8312 - 9.0340 |
| Overall Compliance | 2.5984 | 0.097 | 0.8425 - 8.0131 |
| Female | 0.3414 | 0.086 | 0.0999 - 1.1659 |
| Age 35+ | 1.0789 | 0.893 | 0.3564 - 3.2666 |
| WHO Stage 3 | 1.4222 | 0.662 | 0.2930 - 6.9041 |
| WHO Stage 4 | 0.8582 | 0.89 | 0.0979 - 7.5272 |

WHO-World Health Organization
\*adjusted for age, sex, intervention arm, ART visit compliance, and WHO stage

Most participants returned to the clinic for their appointments (Table 4). More 2wT participants returned on time (81%) compared to SoC participants (74.5%) (p = 0.020). Both late return and no returns were lower among 2wT clients than among clients receiving SoC (18.0% vs 24.1% and 0.97% vs 1.41%, respectively). The Kaplan-Meier curves revealed no difference in retention on ART over time between 2wT and SOC (p=0.4) (S3 Fig 1).

**Table 4:** Association between Intervention Arm and ART visit compliance.

| **ART Visit Compliance** | **SoC (N=640 visits)** |  | **2WT (N=618 visits)** |  |
| --- | --- | --- | --- | --- |
|  | **N** | **%** | **N** | **%** |
| **Returned on time** (within 0-2 days of an appointment) | 477 | 74.5% | 501 | 81.1% |
| **Returned late** (3+ days after an appointment) | 154 | 24.1% | 111 | 18.0% |
| **Never Returned** | 9 | 1.4% | 6 | 1.0% |
| **Pearson Chi2(2) =** | 7.74 |  | Pr= | 0.02 |

## Discussion

Our RCT investigated the impact of 2wT on early retention, compliance to ART appointments, and VLS among new ART initiates within the first six months of ART at LT clinics in Malawi. Although ART retention at 6 months did not differ between groups, 2wT participation was associated with improved clinic visit compliance and higher VLS. Higher VLS may be primarily driven by increased visit compliance. The personalized SMS reminders, motivational messages, and open communication channel provided by 2wT appear to enhance early retention among new ART initiates with phones who elect to participate. These findings align with previous research on SMS interventions with benefits for visit compliance (10,12,14,15,27) and VLS (10,28). These findings suggest several points of consideration.

The RCT results contrast with our earlier quasi-experimental findings that 2wT improved ART outcomes and retention at six and twelve months. This discrepancy may be due to several factors. First, the RCT follow-up was short. It is possible retention gains for 2wT clients might increase over time. Second, the quasi-experimental SoC clients started ART during the COVID-19 pandemic, when retention was lower overall. Third, the quasi-experimental study compared a historical cohort that likely did not receive efficient EC buddy support as the program was in its infancy. Over time, the EC program improved to provide better reminders of on-time visits for SoC clients. Lastly, RCT participants in either intervention or control had verified phone numbers, whereas the comparison group in the quasi-experimental group only had a documented mobile number that was not verified. RCT participants in both groups may simply represent those with higher socio-economic status, making them more likely to comply with visits regardless of intervention (29,30).

Despite mixed results, it appears clear that the 2wT intervention should be expanded to complement existing SoC retention efforts. While 2wT requires both basic literacy and access to a phone, the 2wT benefits for those who are eligible and interested are significant. 2wT also requires fewer financial and human resources, especially at scale (20). If retention is similar between 2wT and SoC, but costs are lower, 2wT scale-up should be recommended for those who are eligible and interested in 2wT support, freeing human and financial resources that could sustain 2wT implementation(19). With the potential workload benefits of 2wT expansion (31,32) combined with the personalized, person-centered approach that aligns well with DSD goals of enhancing client engagement (33), the potential impact of 2wT expansion is high.

### Limitations

First, we could not confirm whether 2wT or SoC received their intended support; fidelity assessment was outside the scope of this small study. Second, VLS data was missing for 123 participants (25%) due to missing samples, a bias that cannot be quantified. Third, participation in the RCT was opt-in, so those who volunteered may have been more open to retention support. Despite these limitations, the results suggest that the lower-cost, lower-intensity 2wT improves VLS and clinic visit compliance, making it a valuable tool for enhancing ART adherence and retention in care.

### Conclusion

2wT improved VLS and appointment compliance within the first six months of ART initiation in a large public ART clinic in Malawi, highlighting 2wT’s potential to complement existing retention support and enhance early ART outcomes. 2wT did not appear to significantly increase ART retention at 6 months, but the impact on ART retention outcomes may be delayed. Nevertheless, if the 2wT approach achieves equal or better visit compliance and VLS with lower-intensity and fewer resources, scaling 2wT alongside continued SoC retention interventions would likely have significant, positive benefits for ART retention and retention support efficiency in high-prevalence, low-resource settings.

## Data Availability

Interested researchers may contact the Human Subjects Division representative for our study at to request access to the datasets. Access to data will be restricted to those who complete data sharing agreements.

## Supporting Information

**S2 Figure 1: CONSORT diagram Study participant flow**

**S2 Figure 1: CONSORT diagram Study participant flow**

**S3 Figure 1: Kaplan-Meier curve of retention on ART among 2wT and SoC clients over time**

## Authors’ contributions

CKK: Writing – original draft.

CKK and RK: Formal Analysis.

CKK, RK, HT, GC, and JH: Investigation and Project Administration.

CKK, JH, JS, AK, DN, PB and AT: Data curation.

CF, WG, MP and HT: Supervision.

HT and CF: Conceptualization.

All authors: Writing – review & editing.

## Acknowledgments

The research reported in this publication was supported by the National Institutes of Health (NIH) under Award Number 3R33TW011658 (CF).

The authors extend their gratitude to the Lighthouse Trust retention program and research department for their collaboration in implementing the 2wT intervention.

## Notes

### Competing Interest Statement

The authors have declared no competing interest.

### Clinical Trial

The study was registered with the Clinical Trial Registry on 9/6/2022 (NCT05531448).

### Funding Statement

Yes

